# Sixty-day mortality among 520 Italian hospitalized COVID-19 patients according to the adopted ventilatory strategy in the context of an integrated multidisciplinary clinical organization: a population-based cohort study

**DOI:** 10.1101/2020.08.13.20174615

**Authors:** Antonella Potalivo, Jonathan Montomoli, Francesca Facondini, Gianfranco Sanson, Luigi Arcangelo Lazzari Agli, Tiziana Perin, Francesco Cristini, Enrico Cavagna, Raffaella De Giovanni, Carlo Biagetti, Ilaria Panzini, Cinzia Ravaioli, Maddalena Bitondo, Daniela Guerra, Giovanni Giuliani, Elena Mosconi, Sonia Guarino, Elisa Marchionni, Gianfilippo Gangitano, Ilaria Valentini, Luca Giampaolo, Francesco Muratore, Giuseppe Nardi

**Affiliations:** Infermi Hospital, Rimini (Italy); Dept. of Medicine, Surgery and Health Sciences, University of Trieste (Italy); AUSL della Romagna Health Care Service, Ravenna (Italy)

## Abstract

**Background:** Among COVID-19 patients, the decision of which ventilation strategy to adopt is crucial and not guided by existing outcome evidence. We described the clinical characteristics and outcomes of hospitalized COVID-19 patients according to the adopted respiratory strategy.

**Methods:** Population-based cohort study including all COVID-19 patients (26/02/2020-18/04/2020) within Rimini Italian province. Hospitalized patients were classified according to the maximum level of respiratory support: oxygen supplementation (group Oxygen), NIV (group NIV-only), IMV (group IMV-only), and IMV after a NIV trial (group IMV-after-NIV). Sixty-day mortality risk was estimated with a Cox proportional hazard analysis adjusted by age, sex, and administration of steroids, canakinumab, and tocilizumab.

**Findings:** We identified 1,424 symptomatic patients: 520 (36.5%) were hospitalized, the remaining 904 (63.5%) were treated at home with no 60-days death. According to the respiratory support, 408 (78.5%) were assigned to Oxygen, 46 (8.8%) to NIV-only, 25 (4.8%) to IMV-after-NIV, and 41 (7.9%) to IMV-only groups. There was no significant difference in the P/F at IMV inception among IMV-after-NIV and IMV-only groups (*p*=0.9). Overall 60-day mortality was 24.2% (Oxygen: 23.0%; NIV-only: 19.6%; IMV-after-NIV: 32.0%; IMV-only: 36.6%; *p* = 0.165). Compared with Oxygen group, the 60-day mortality risk significantly increased for IMV-after-NIV (HR 2.776; *p*=0.024) and IMV-only group (HR 2.966; *p*=0.001).

**Conclusions:** This study provides a population-based figure of the impact of the COVID-19 epidemic. A similar 60-days mortality risk was found for patients undergoing immediate IMV and those intubated after a NIV trial. Many patients had a favorable outcome after prolonged IMV.

## Introduction

More than 650,000 people died worldwide from the coronavirus-disease 2019 (COVID-19) caused by the severe acute respiratory syndrome coronavirus 2 (SARS-CoV-2). The percentage of COVID-19 patients requiring non-invasive (NIV) or invasive mechanical ventilation (IMV) is unclear and strongly affected by the hospital organization and the availability of resources. Three studies from China, US, and Gemany reported the use of IMV among hospitalized COVID-19 patients to be 2.3%, 12%, and 17%, respectively.^1–3^ ICU-mortality among mechanically ventilated COVID-19 patients varies from 25% to 97%.^2-10^ Such wide variations may have different explanations. First, few studies included all patients hospitalized for COVID-19 able to provide a complete overview of characteristics and outcomes of COVID-19 patients that required hospitalization. Second, information regarding the respiratory management of COVID-19 patients has been mainly described in the setting of the intensive care units (ICUs).^4,8,9,11^ Thus, the number of hospitalized COVID-19 patients treated with oxygen supplementation and NIV has been markedly underreported leading to inaccurate information regarding the overall use of the different respiratory supports and outcomes. Third, most previous reports included a percentage of patients still admitted at the ICU at the end of the follow-up ranging from 2.3%^8^ to 71%^2^ and this may have led to different degrees of inaccuracy in estimating ICU-mortality. Finally, ICU beds and human resources availability is likely to vary across different areas and it has not been generally described in clinical studies, affecting generalizability of the results.^12^

The Province of Rimini in Northern Italy was one of the areas more severely affected by the COVID-19 outbreak. On March 7^th^, Rimini province was declared “Red Zone” due to the high number of infected people and, therefore, it was isolated with no possibility of entry or exit. Using clinical and demographics information routinely collected in a unique database including all residents in the entire province, we performed the present population-based cohort study with the following aims: 1) to describe the characteristics of hospitalized COVID-19 patients, 2) to examine patient outcomes overall and stratified by the adopted respiratory support, 3) to describe the organization of local healthcare system.

## Methods

### Setting

COVID-19 patients admitted to the Rimini hospital were retrieved in the present observational population-based cohort study, following the Strengthening the Reporting of Observational Studies in Epidemiology (STROBE) reporting guideline. The Italian National Public Healthcare System provides homogeneous and free access to any level of appropriate treatment to all the people including irregular immigrants through the hospital network, family doctors, and District Health Systems. The province of Rimini consists of approximately 340,000 inhabitants and is served by a network of five public hospitals, with Rimini Hospital being the largest and providing up to 600 beds.

Since the beginning of the outbreak and for the entire duration of this study, Rimini Hospital was identified as reference hospital for all COVID-19 positive or suspected patients. With the increase of admitted patients, 340 hospital beds - including 80 in two newly opened wards- were progressively dedicated to infected subjects and 28 negative pressure rooms –eight set up at Emergency Department (ED) and 20 already available in the Infectious Diseases ward– were dedicated to patients requiring NIV. During the first decade of March, the number of ICU beds was progressively increased from 20 to 53, of which 48 dedicated to COVID-19 patients and five to non-COVID-19 patients. Non-COVID-19 patients exceeding local availability were transferred to another nearby hospital equipped with 10 ICU beds.

### Study population

All patients evaluated at one of the five EDs of the province from February, 26 to April, 18 2020 presenting symptoms suspicious for COVID-19 infection (i.e. fever and/or respiratory symptoms) and tested for the SARS-CoV-2 (real-time RT-PCR)^13^ were considered for inclusion. Patients with positive swab, as well those with chest X-ray or CT scan evidence of COVID-19-related pneumonia despite a negative swab were included. The more stable patients were discharged home and entrusted to the primary care, while patients identified to be at high risk for complications based on symptoms severity and associated comorbidities were admitted at the hospital and represented the study population. All included patients with a first negative swab had at least a positive subsequent swab during the same hospitalization except those patients that died at the ED before a second swab.

### Organization

A daily meeting –always attended by the heads of the Emergency, Infectious Disease, Pneumology, Radiology, and Intensive Care Departments– was planned regardless of the holidays or Sundays respecting airborne and contact transmission precautions, with the aim to ensure a homogeneous and standardized treatment to all COVID-19 patients. Every day the relevant clinical information (e.g., comorbidities, respiratory status, medical treatments, and active clinical conditions) of the critically ill cases were updated and the overall therapeutic stewardship to be adopted (e.g., off-label medications, change in the respiratory support) was collegially discussed and agreed. Moreover, for each patient the appropriate treatment to adopt in the event of worsening conditions was planned, taking into account the limitation of the available resources. All decisions were recorded and promptly communicated to physicians and nurses working at all COVID-19 wards, comprising the ICU.

### Respiratory support

Oxygen administration via ventimask or mask with reservoir was considered the standard of care for moderate/severe patients, while NIV (comprising continuous positive airway pressure, CPAP) and IMV following tracheal intubation were chosen for the most critical cases. To avoid airborne viral transmission, high flow nasal cannulae were used only for respiratory weaning in patients become negative and helmet was identified as the only interface to be used for NIV. Conditions leading to the decision to start IMV were: respiratory fatigue, new hemodynamic instability, or worsening of gas exchange notwithstanding oxygen/NIV, also considering prognostic criteria and resources availability (e.g., ICU beds and mechanical ventilators). According to the level of the respiratory support, the study population was divided in the following subgroups: Oxygen (patients receiving no more than oxygen supplementation); NIV-only (patients receiving no more than NIV); IMV-after-NIV (patients undergoing IMV after a failed NIV trial); IMV-only (patients starting IMV at hospital admission or after a trial with oxygen).

### Data sources, follow-up and outcomes

For the whole population, demographic data were retrieved by an administrative and clinical database (Maria DB, Log 80, Forli-Italy) and information about administered off-label medications related to the COVID-19 treatment (i.e. hydroxichloroquine, antivirals, steroids, canakinumab, and tocilizumab) were collected by clinical documentation. For patients treated with NIV and/or IMV the Charlson comorbidity index was computed,^14^ and the SpO_2_ at the hospital admission, the PaO_2_/FiO_2_ (P/F) ratio at the inception of NIV and IMV, and the duration of ventilatory supports were collected. For those patients the extent of lung damage was estimated from the chest radiogram using the Brixia score: each lung was transversally divided into three sectors and to each obtained sector a score ranging from zero (no alteration) to three (interstitial-alveolar infiltrates) was assigned (total score range: 0–18).^15,16^

For patients admitted to the ICU (i.e., IMV-after-NIV and IMV-only), the Simplified Acute Physiology Score II (SAPS 2)^17^ and the Sequential Organ Failure Assessment (SOFA) score^18^ at ICU admission were computed and the possible implementation of tracheostomy and renal replacement therapy were documented.

All patients were followed up to 60 days from hospital admission. The condition of being dead or alive at this time constituted the main study endpoint. Accordingly, data collection was concluded on June 18, 2020 to ensure at least 60 days of observation to the patients included last.

### Ethical aspects

The investigation conforms with the principles outlined in the Declaration of Helsinki. The AUSL della Romagna Institutional Review Board approved the project (registration number NCT04348448) with a waiver of informed consent. No additional procedure or investigation potentially related to the study was requested or provided.

### Statistics

Continuous variables were described as mean ± standard deviation. The differences between the means were analysed by a paired or unpaired Student’s t-test, as appropriate, after considering whether the subgroups had equal variance using Levene’s test. One-way analysis of variance (ANOVA) was applied for all comparisons between the subgroups. The nominal variables were presented as numbers and percentages and compared either through χ test or Fisher’s exact test, as appropriate.

The ability of the P/F measured before a NIV trial to predict NIV failure (i.e., death or need of IMV) was tested by calculating the area under the receiver operating characteristics curve (AUC). The maximum Youden index (*J*) was considered as the optimal P/F cut-off value. Sixty-day mortality was computed using Kaplan-Meier technique overall and among groups. The Mantel-Cox log-rank test was adopted to assess differences among the survival rates. A multivariate Cox proportional hazard analysis was used to estimate 60-day mortality risk among the study groups in comparison to the Oxygen group (reference group), adjusted by age, sex, and administration of steroids, canakinumab, and tocilizumab. The results were presented as a proportional hazard ratio (HR) with 95% CI and adjusted cumulative survival curves. Finally, in order to examine the potential impact of survival bias among patients treated with IMV, especially among patients intubated after NIV (a patient had to survive until endotracheal intubation), we computed the adjusted HRs for 10-day and 11 to 60-day mortality, separately, as sensitivity analysis. Moreover, minimally to fully adjusted HRs were reported in the Supplemenatry material.

For all tests, statistical significance was set at an alpha level of *p* = 0.05. All statistical analyses were performed using the software IBM SPSS Statistics, version 24.0 (Armonk, NY, US: IBM Corp.).

## Results

During the study period, 1,424 symptomatic patients were evaluated at the EDs in the province and had a positive swab and/or chest imaging suspicious for the COVID-19. Nine-hundred and four (63.5%) were treated at home without further ED accesses, while the remaining 520 (36.5%) were hospitalized and constituted the study population (males 350, 67.3%; mean age 70.7 ± 14.1, range 19-98). Among hospitalized patients, 440 (84.6%) were treated with oxygen supplementation at the time of hospital admission, while the remaining received either NIV or IMV. During the subsequent days, 57 patients (11.0%) required an upgrade of the ventilatory support. Almost all patients were treated with hydroxichloroquine and antiviral drugs. Table 1 describes the demographic characteristics and the administered medications of the study population overall and by groups. Figure 1 shows a complete synthesis of the respiratory support adopted and the 60-days mortality for each group.

**Table 1.**
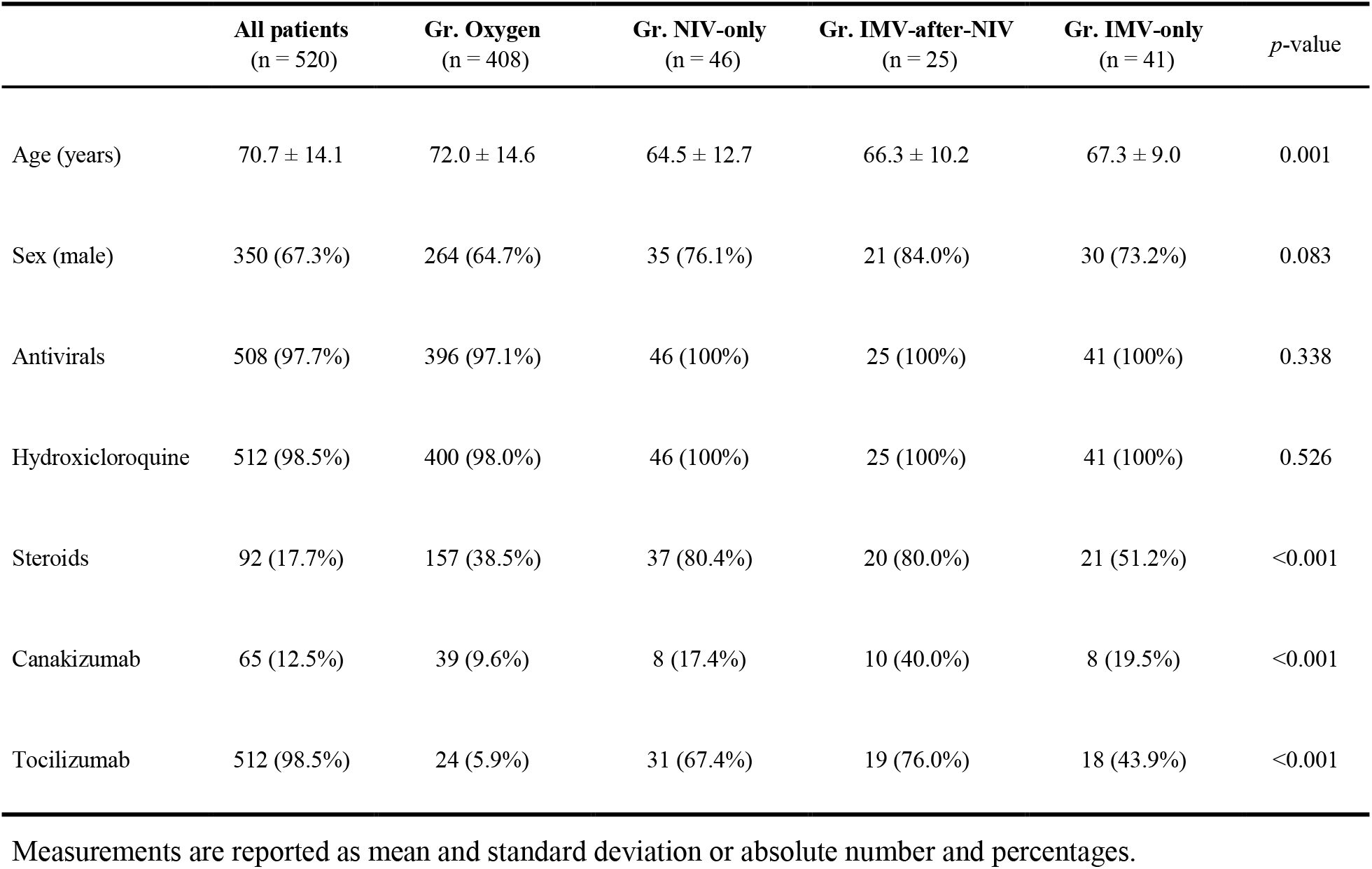
Comparison of demographic characteristics and administered medications among the study subgroups.

**Figure 1.**
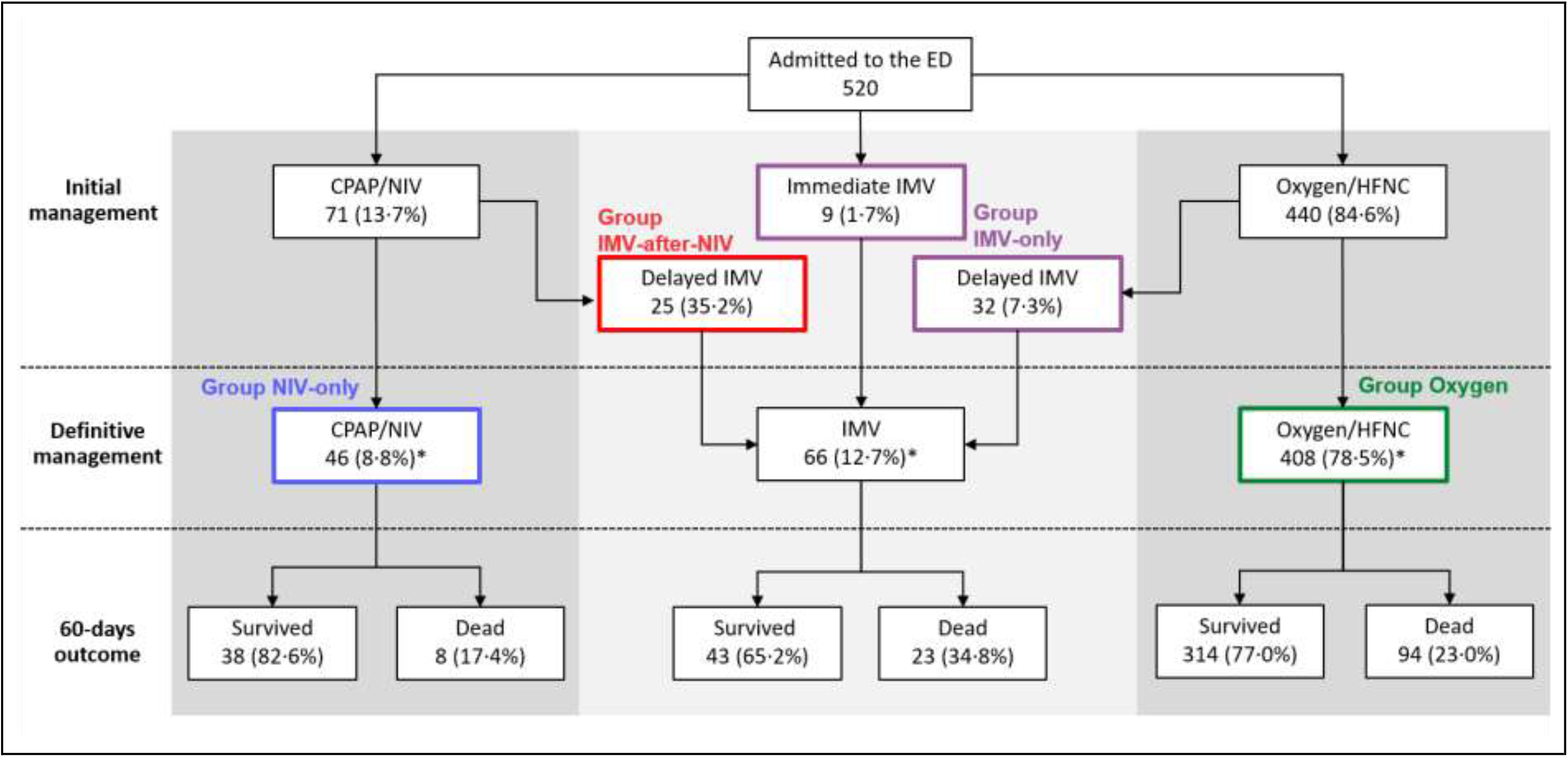
Flow chart synthesizing the clinical pathways of the COVID-19 patients, the respiratory support provided at each step of their hospital stay and their 60-days mortality. Percentages are referred to the previous level in the flow-chart, unless otherwise indicated. *: with respect to total hospital admissions. CPAP: Continuous Positive Airway Pressure. NIV: Non-invasive ventilation. IMV: Invasive mechanical ventilation. ED: emergency department

### Ventilatory support

The main clinical characteristics of the 112 (21.5%) patients who received any ventilatory support (NIV and/or IMV, mean age 66.9 ± 9.4 years) are reported in Table 2. Among those, 71 (63.4%) patients received at least one trial of NIV (mean age 65.1 ± 11.8) with a mean duration of 3.8 ± 2.2 days (range 1-10, Figure 2a) and a mean P/F ratio of 105.9 ± 40.6 at the beginning of NIV. Thirty-eight (53.5%) of the patients treated with NIV improved and were transferred to a COVID-19 ward, while 25 (35.2%) were intubated and admitted to the ICU to undergo IMV. The P/F ratio before starting NIV differed significantly between patients with a successful trial and those that failed (successful: 119.4 ± 46.2; failing: 92.1 ± 23.8; *p* = 0.003). For patients who needed IMV a statistically significant but clinically irrelevant improvement in P/F ratio was documented (before NIV: 93.1± 23.8; before IMV: 113.8 ± 41.5; *p* = 0.041). The remaining eight patients (11.3%) died without being intubated (P/F ratio before NIV 85.3 ± 36.5). The length of NIV did not differ among patients with successful or failing trial (Table 2). The ability of the P/F ratio obtained before NIV to predict the failure of a NIV trial showed an AUC of 0.71 (95% CI 0.59-0.83; p-value 0.002) and provided a best cut-off of 115.5 (sensitivity 52.6%, specificity 81.8%; *J* 0.353).

**Table 2.** Characteristics of patients receiving either invasive or non-invasive mechanical ventilation.

| Variable | Group NIV-only<br>n; mean ± SD | Group IMV-after-NIV<br>n; mean ± SD | Group IMV-only<br>n; mean ± SD | *p*-value |
| --- | --- | --- | --- | --- |
| Charlson comorbidity index | 44; 2.5 ± 1.6 | 23; 2.3 ± 1.1 | 40; 2.6 ± 2.3 | 0.778 |
| SpO<sub>2</sub> on H admission | 42; 91.8 ± 4.7 | 25; 87.4 ± 8.0 | 40; 89.6 ± 6.2 | 0.019 |
| Brixia score before NIV | 43; 12.2 ± 2.6 | 22; 10.9 ± 3.7 | .. | 0.115 |
| Brixia score before IMV | .. | 23; 13.8 ± 2.8 | 39; 13.8 ± 2.5 | 0.986 |
| H-admission to NIV (days) | 46; 2.2 ± 2.6 | 25; 2.1 ± 2.8 | .. | 0.884 |
| P/F before NIV | 46; 113.4 ± 46.2 | 25; 92.1 ± 23.8 | .. | 0.013 |
| Length of NIV (days) | 46; 3.9 ± 2.0 | 25; 3.6 ± 2.6 | .. | 0.546 |
| H-admission to IMV (days) | .. | 25; 7.0 ± 6.0 | 41; 3.8 ± 5.5 | 0.027 |
| P/F before IMV | .. | 24; 113.8 ± 41.5 | 36; 112.5 ± 50.4 | 0.919 |
| SAPS II on ICU admission | .. | 23; 41.6 ± 12.8 | 40; 50.8 ± 19.5 | 0.028 |
| SOFA score on ICU admission | .. | 23; 6.9 ± 4.0 | 40; 9.0 ± 3.6 | 0.033 |
| Length of IMV (days) | .. | 25; 19.5 ± 15.2 | 41; 24.5 ± 20.9 | 0.307 |
| IMV to tracheostomy (days) | .. | 17; 9.9 ± 4.1 | 29; 9.8 ± 3.1 | 0.909 |
| Length of stay in ICU (days) | .. | 24; 19.7 ± 14.9 | 40; 26.0 ± 22.2 | 0.176 |

SD: standard deviation. NIV: non-invasive ventilation; IMV: invasive mechanical ventilation; H: hospital; SpO<sub>2</sub>: peripheral oxygen saturation; P/F: PaO<sub>2</sub>/FiO<sub>2</sub> ratio; SOFA: sequential organ failure assessment; SAPS: simplified acute physiology score; ICU: intensive care unit.

**Figure 2.**
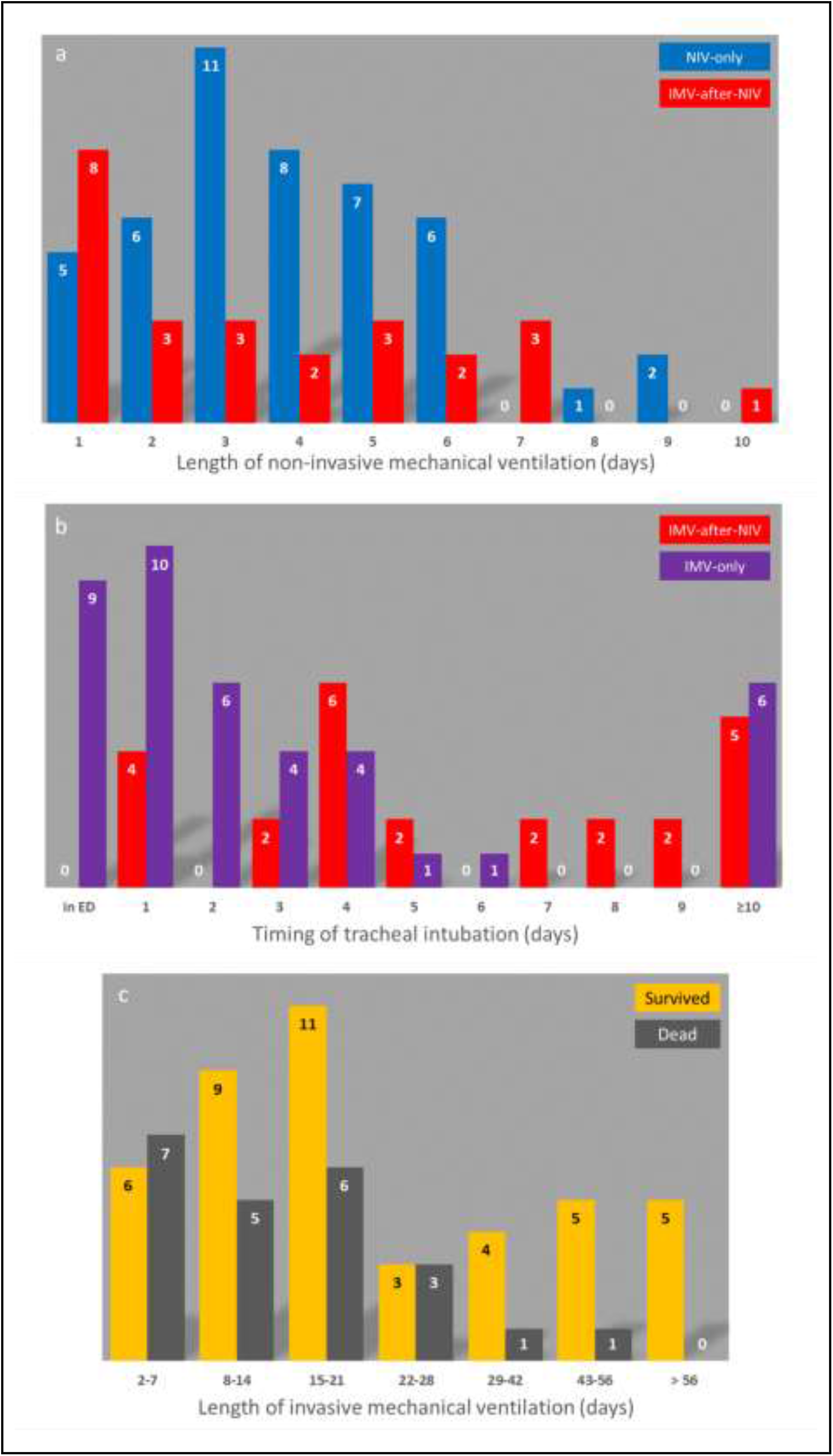
**a)** Duration of continuous non-invasive ventilation (NIV) for patients who received at least one trial of NIV (n = 71). **b)** Interval between hospital admission and the onset of invasive mechanical ventilation (IMV) for patients admitted to the intensive care unit (n = 66). **c)** Length of IMV in patients who survived or dead at 60- days follow-up.

Overall, 66 patients (Groups IMV-after-NIV and IMV-only, mean age 66.9 ± 9.4 years) were admitted to the ICU and treated with IMV (Figure 1). The mean interval between hospital admission and the onset of IMV was 5.0 ± 5.9 days (range 0-26) (Figure 2b). Eleven (15.2%) patients were intubated after 10 or more days from hospital admission (P/F before intubation: 113.0 ± 46.7). No difference in P/F ratio at the time of the definitive ventilation support was found among the three study groups *(p* = 0.993).

Patients who failed the NIV trial (n = 21) had a Brixia score of 10.6 ± 3.4 before NIV which worsened to 13.4 ± 2.5 (p = 0.002) before IMV. Similar Brixia score was obtain at the time of endotracheal intubation among patients that failed a NIV trial and among patients that were treated with IMV without a NIV trial (Table 2).

Among the 66 patients admitted to the ICU, mean duration of IMV was 22.6 ± 19.0 days (range 2–87). Forty-six ICU patients (69.7%) received percutaneous tracheostomy (time from intubation: 9.8 ± 3.5 days). Renal replacement therapy was performed in 19 (28.8%) patients, with a mean duration of 21.7 ± 18.7 days.

### Outcome

None of the 904 patients treated at home died during the follow-up. The overall 60-days mortality for hospitalized patients was 24.2% (n = 126), and was 23.0% (n = 94), 19.6% (n = 9), 32.0% (n = 8) and 36.6% (n = 15) for Group Oxygen, NIV- only, IMV-after-NIV, and IMV-only, respectively (*p* = 0.165). Mortality among the 112 patients receiving any ventilatory support (NIV and/or IMV) was 27.7%. No between-groups difference in mortality was found by comparing the crude Kaplan-Meier curves (Log-rank test: *p* = 0.343, Figure 3a). Age was a risk factor for death in Groups Oxygen and NIV-only but not for patients undergoing IMV. The relationships between some characteristics of the study groups and mortality are described in Table 3.

**Table 3.** Comparison of mortality rate among study subgroups

|  | Group Oxygen |  |  | Group NIV-only |  |  | Group IMV-after-NIV |  |  | Group IMV-only |  |  |
| --- | --- | --- | --- | --- | --- | --- | --- | --- | --- | --- | --- | --- |
|  | Survived<br>(n = 314) | Dead<br>(n = 94) | *p*-value | Survived<br>(n = 38) | Dead<br>(n = 8) | *p*-value | Survived<br>(n = 17) | Dead<br>(n = 8) | *p*-value | Survived<br>(n = 26) | Dead<br>(n = 15) | *p*-value |
| Age (years) | 68.8 ± 14.6 | 82.8 ± 8.1 | <0.001 | 61.7 ± 12.2 | 75.9 ± 7.1 | 0.002 | 64.9 ± 11.3 | 69.2 ± 6.8 | 0.332 | 65.8 ± 9.2 | 69.7 ± 8.4 | 0.185 |
| Sex (male) | 197 (62.7%) | 67 (71.3%) | 0.129 | 28 (75.7%) | 7 (77.8%) | 1.000 | 16 (94.1%) | 5 (62.5%) | 0.081 | 18 (69.2%) | 12 (80.0%) | 0.716 |
| Steroids | 122 (38.9%) | 35 (37.2%) | 0.777 | 33 (89.2%) | 4 (44.4%) | 0.002 | 12 (70.6%) | 8 (100%) | 0.140 | 15 (57.7%) | 6 (40.0%) | 0.275 |
| Immunotherapy | 44 (14.0%) | 15 (16.0%) | 0.638 | 33 (89.2%) | 4 (44.4%) | 0.008 | 15 (88.2%) | 8 (100%) | 1.000 | 16 (61.5%) | 7 (46.7%) | 0.355 |
| Tocilizumab | 17 (5.4%) | 7 (7.4%) | 0.462 | 27 (73.0%) | 4 (44.4%) | 0.127 | 12 (70.6%) | 7 (87.5%) | 0.624 | 11 (42.3%) | 7 (46.7%) | 0.786 |
| Canakinumab | 31 (9.9%) | 8 (8.5%) | 0.694 | 8 (21.6%) | 0 (0.0%) | 0.324 | 9 (52.9%) | 1 (12.5%) | 0.088 | 8 (30.8%) | 0 (0.0%) | 0.018 |
| Hyd.chloroquine | 310 (98.7%) | 90 (95.7%) | 0.067 | 37 (100%) | 9 (100%) | .. | 17 (100%) | 8 (100%) | .. | 26 (100%) | 15 (100%) | .. |
| Antivirals | 307 (97.8%) | 89 (94.7%) | 0.120 | 37 (100%) | 9 (100%) | .. | 17 (100%) | 8 (100%) | .. | 26 (100%) | 15 (100%) | .. |

**Figure 3.**
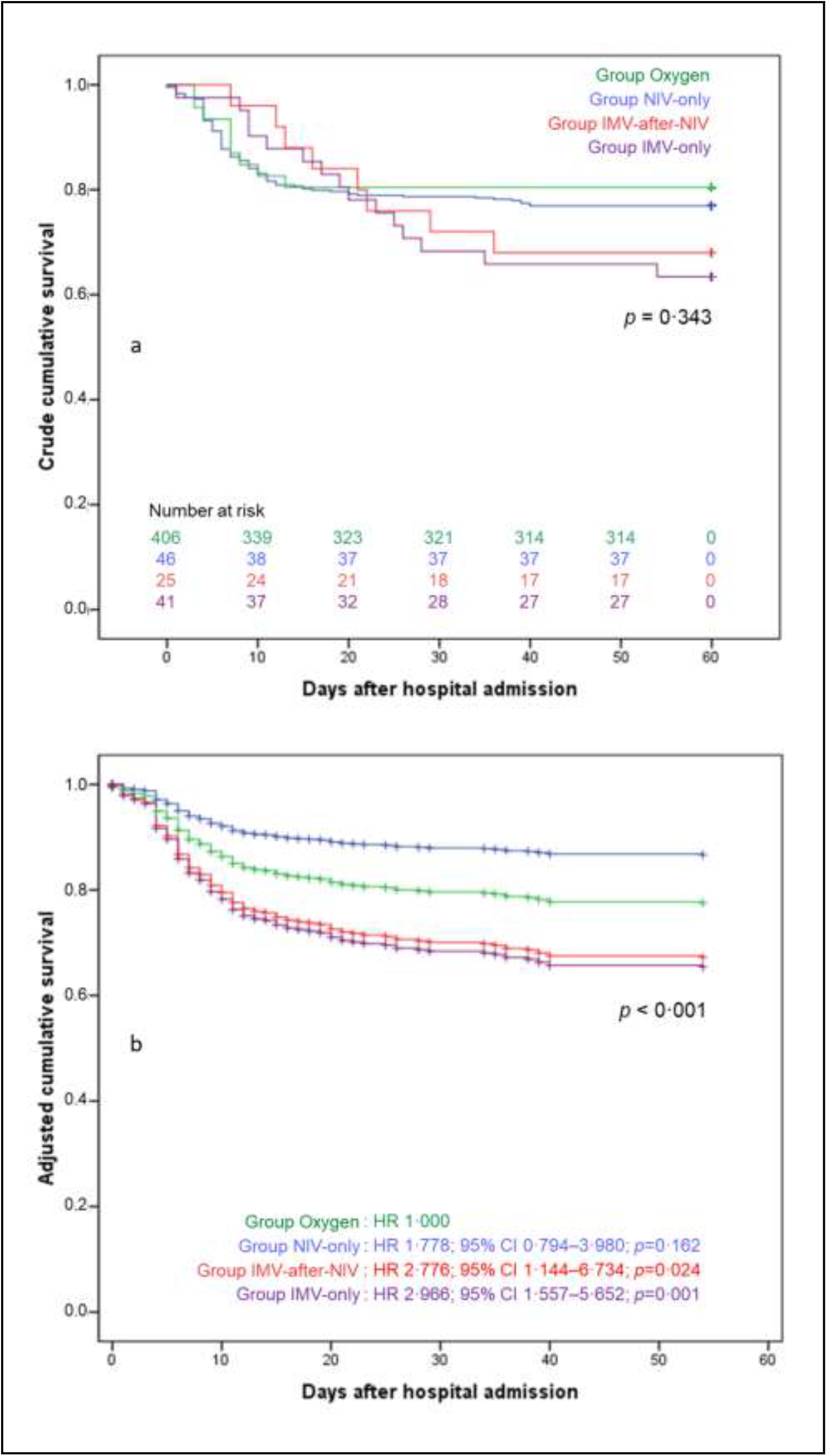
Crude (**a**) and adjusted (**b**) Kaplan-Meier curves for the risk of 60-day mortality in patients belonging to the study groups according to provided respiratory support.

The mean duration of IMV was 26.5 ± 21.1 days for the 43 patients alive at the end of the follow-up (range: 6-70) and 15.3 ± 11.5 days for the 23 non-survivors (range: 2-51). Among the 43 survived, 14 patients (32.6%) underwent IMV for more than four weeks (Figure 2c). Two patients (one belonging to the IMV-after-NIV group and one to the IMV-only group) were still on IMV at the end of follow-up, with an IMV duration of 70 and 63 days, respectively.

Compared with Oxygen group, the multivariate Cox regression analysis showed a 60-day mortality risk progressively increasing among the other groups being statistically significant for the IMV-after-NIV group (HR 2.776; 95% CI 1.144–6.734; *p* = 0.024) and the IMV-only group (HR 2.966; 95% CI 1.557–5.652; *p*=0.001) but not for Group NIV-only (HR 1.778; 95% CI 0.794–3.980; *p* = 0.162) (Figure 3b). Among the explored covariates, older age (HR 1.097; 95% CI 1.073–1.120; p < 0.001) and male sex (HR 1.597; 95% CI 1.072–2.379; *p* = 0.021) showed a statistically significant association with the mortality. As expected, the sensitivity analysis showed a higher risk for 10-day mortality in the only-NIV group but not in patients receiving IMV compared to patients treated with oxygen, while similar results to the main analysis were provided for the 11 to 60-day mortality (Table 4 and Figure 4).

**Table 4.** Results of multivariate Cox proportional hazard analysis of mortality at different time interval from hospital admission for patients undergoing to different breathing support modalities.

|  | **Breathing support** | **60-days mortality**<br>HR (95% CI); *p*-value | **10-days mortality**<br>HR (95% CI); *p*-value | **11- to 60-days mortality**<br>HR (95% CI); *p*-value |
| --- | --- | --- | --- | --- |
| Model #0 | Oxygen* | 1.000 | 1.000 | 1.000 |
|  | NIV-only | 0.839 (0.424-1.663); 0.616 | 1.017 (0.489-2.115); 0.963 | 0.350 (0.047-2.583); 0.303 |
|  | IMV-after-NIV | 1.307 (0.635-2.690); 0.468 | 0.220 (0.031-1.583); 0.133 | 4.408 (1.906-10.197); 0.001 |
|  | IMV-only | 1.538 (0.892-2.652); 0.122 | 0.547 (0.200-1.500); 0.241 | 4.418 (2.173-8.984); <0.001 |
| Model #1 | Oxygen* | 1.000 | 1.000 | 1.000 |
|  | NIV-only | 1.890 (0.925-3.859); 0.081 | 2.345 (1.084-5.072); 0.030 | 0.707 (0.093-5.392); 0.738 |
|  | IMV-after-NIV | 2.847 (1.326-6.111); 0.007 | 0.530 (0.072-3.919); 0.534 | 7.626 (3.059-19.012); <0.001 |
|  | IMV-only | 3.074 (1.708-5.532); <0.001 | 1.181 (0.415-3.361); 0.755 | 7.312 (3.337-16.021); <0.001 |
| Model #3 | Oxygen* | 1.000 | 1.000 | 1.000 |
|  | NIV-only | 1.793 (0.878-3.660); 0.109 | 2.202 (1.019-4.759); 0.045 | 0.691 (0.090-5.271); 0.721 |
|  | IMV-after-NIV | 2.689 (1.250-5.785); 0.011 | 0.499 (0.067-3.692); 0.496 | 7.376 (2.938-18.515); <0.001 |
|  | IMV-only | 3.037 (1.687-5.468); <0.001 | 1.172 (0.412-3.339); 0.766 | 7.224 (3.295-15.838); <0.001 |
| Model #3 | Oxygen* | 1.000 | 1.000 | 1.000 |
|  | NIV-only | 1.910 (0.926-3.944); 0.080 | 2.650 (1.216-5.772); 0.014 | 0.533 (0.069-4.120); 0.546 |
|  | IMV-after-NIV | 2.869 (1.319-6.239); 0.008 | 0.629 (0.084-4.686); 0.651 | 6.012 (2.344-15.414); <0.001 |
|  | IMV-only | 3.071 (1.706-5.529); <0.001 | 1.182 (0.416-3.357); 0.754 | 6.623 (3.008-14.583); <0.001 |
| Model #4 | Oxygen* | 1.000 | 1.000 | 1.000 |
|  | NIV-only | 1.936 (0.937-4.001); 0.074 | 2.663 (1.222-5.803); 0.014 | 0.539 (0.069-4.180); 0.554 |
|  | IMV-after-NIV | 3.075 (1.405-6.730); 0.005 | 0.655 (0.087-4.933); 0.682 | 6.542 (2.549-16.792); <0.001 |
|  | IMV-only | 3.160 (1.753-5.695); <0.001 | 1.199 (0.421-3.412); 0.734 | 6.888 (3.121-15.203); <0.001 |
| Model #5 | Oxygen* | 1.000 | 1.000 | 1.000 |
|  | NIV-only | 1.778 (0.794-3.980); 0.162 | 3.361 (1.406-8.033); 0.006 | 0.320 (0.038-2.666); 0.292 |
|  | IMV-after-NIV | 2.776 (1.144-6.734); 0.024 | 0.897 (0.111-7.277); 0.919 | 3.689 (1.223-11.133); 0.021 |
|  | IMV-only | 2.966 (1.557-5.652); 0.001 | 1.433 (0.484-4.245); 0.516 | 4.744 (1.918-11.734); 0.001 |

HR: hazard ratio. CI: confidence interval. Model #0: unadjusted; Model #1: adjusted by age; Model #2: adjusted by age and sex; Model #3: adjusted by age, sex, and administration of steroids; Model #4: adjusted by age, sex, and administration of steroids, and canakinumab; Model #5: adjusted by age, sex, and administration of steroids, canakinumab, and tocilizumab. \*: reference

**Figure 4.**
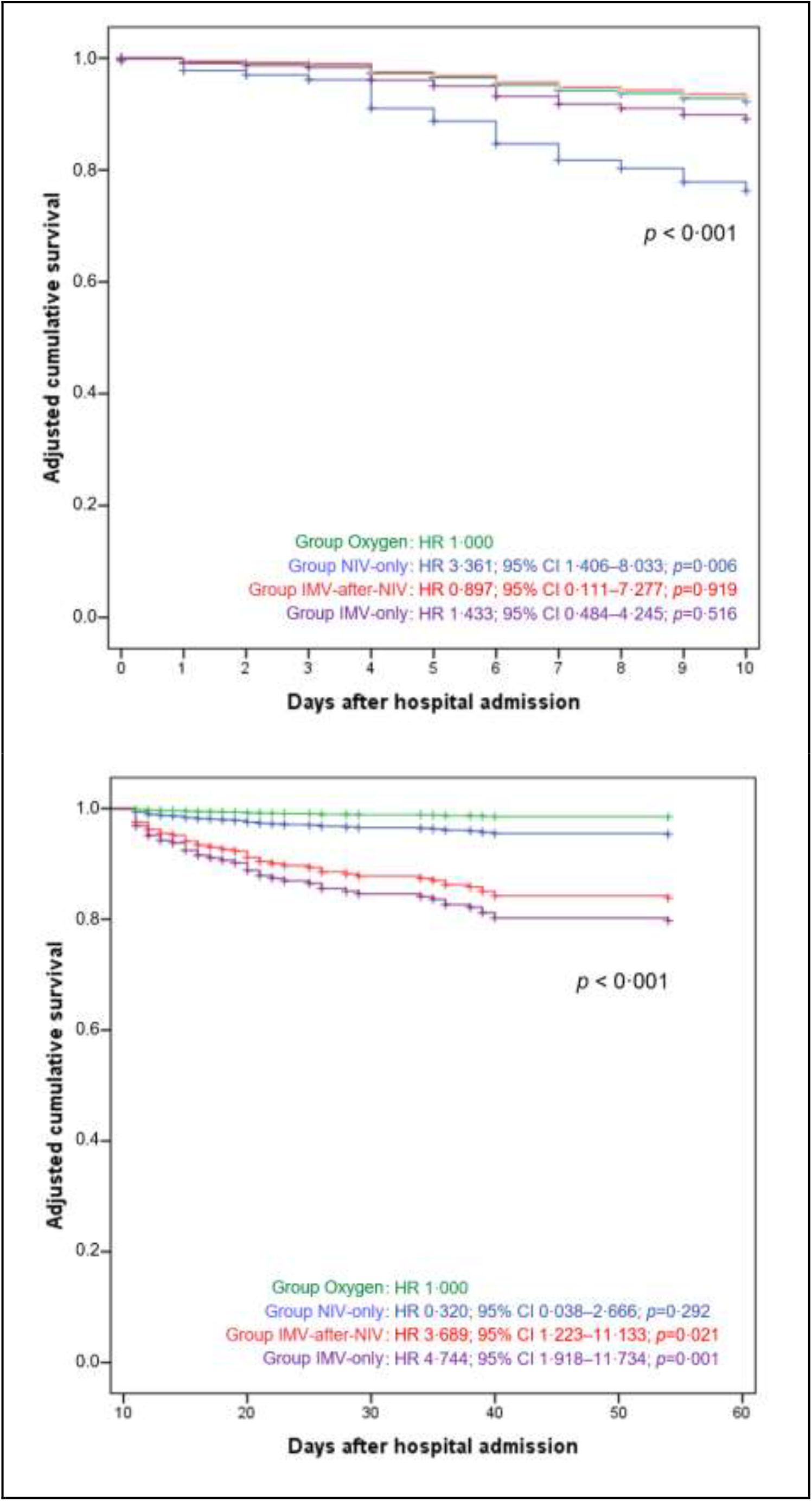
Adjusted Kaplan-Meier curves for the risk of 10- and 11 to 60-day mortality in patients belonging to the study groups according to provided respiratory support.

## Discussion

### Mortality according to breathing support

The present study showed how an integrated multidisciplinary clinical organization allowed to optimize the allocation of the available resources among 520 hospitalized COVID-19 patients. The overall 60-day mortality was 24.2%. Approximately 21% of patients were mechanically ventilated, with a mortality ranging from 19.6% in patients treated with NIV-only to 36.6% in patients undergoing IMV without a NIV trial. To our knowledge, this is the first study reporting 60-day mortality in a cohort of hospitalized patients diagnosed with COVID-19 overall and according to all adopted ventilatory strategies. A recent Chinese multicentric study^19^ enrolling ICU 258 patients reported an overall 60-day mortality of 64.3%, with 19 patients deceased within 48h after ICU admission. Among 165 mechanically ventilated patients, the 60-day mortality was 83%, 56%, and 94% for those treated with IMV, with NIV, and receiving both treatments, respectively. Median P/F ratio in the Chinese population was 91 (IQR 67-134) and SOFA score 6 (IQR 5-7). It should be noted that in our study we documented for patients treated with NIV and/or IMV –despite a similar P/F ratio (median 98.0; IQR 84.0-124.5) and a higher SOFA score (median 8; IQR 6-10)– a considerably lower 60-day mortality rate, overall and in individual groups. A study from six COVID ICUs from US^4^ enrolled 217 patients, 165 of which (76.0%) received IMV, with a median IMV length of 9 days (IQR 4–13). Among IMV subjects, ICU mortality was 33.9% without any difference in IMV days between deceased and survived. At the end of follow-up (median observation time 15 [IQR: 9-24] days), hospital mortality was 35.7% (59/165), being 8 patients still in ICU on IMV. However, no information was provided neither about the adopted respiratory support, nor the outcome, for patients not undergoing IMV. Despite mortality reported by Auld et al. is similar to the mortality reported in our study in the IMV-only group, they reported a much shorter follow-up (maximum follow-up 60 days) and 4.8% of patients were still admitted to the ICU. Therefore, 60-day mortality among those patients is likely to be higher that the reported hospital mortality. Recently, a nationwide study from Germany including more than 10,000 hospitalized COVID-19 patients was published. Interestingly, despite the Germany health-care system has not been overwhelmed by the pandemic the reported inhospital mortality was markedly higher among patients treated with NIV (45% in patients with successfully NIV, 50% in patients that failed NIV) and IMV (53% in patients treated with IMV), while it was lower in patients without mechanical ventilation (16%).^3^

Two further studies considering ICU patients reported information about all adopted breathing support strategies. An Italian multicentric study^8^ enrolling a cohort of 3,355 critically ill patients (median follow-up: 69 days; ventilatory support: IMV 87%, NIV 10%, CPAP/oxygen 2.3%) reported a mortality rate at the censoring (median observation time 70 [range, 38-112] days) of 17%, 36%, and 52% among patients treated with oxygen, NIV and IMV, respectively. Accordingly, NIV and IMV were associated with an increased risk of death compared with patients treated only with oxygen (HR 2.36, 95% CI 1.33-4.17 and HR 3.77, 95% CI 2.19-6.51, respectively). Again, mortality reported in the present study is likely to underestimate 60-day mortality and mortality in the NIV and IMV groups is higher than the reported in our study population. It should be noted that in all above cited studies the reported mortality for patients treated with standard oxygen and NIV was clearly conditioned by the reduced size of these subgroups, related to the study settings limited to ICU.

Compared with the previously reported literature, mortality reported in our study is generally lower. The centralized multidisciplinary approach adopted at Rimini hospital may partially explain the difference with the existing literature. The sensitivity analysis and the differences between the crude and the adjusted HRs reported in the Figure 4 and Table 4 may help with the interpretation of the study findings. Interestingly, in the present investigation, mortality was higher in the oxygen and NIV-only groups in the first 10 days compared to the other two groups. This is partially explained by the survival bias among the mechanically ventilated patients, especially among patients that were intubated after failing a NIV trial. Indeed, although HRs for the 10-day mortality risk did not reach the statistical significance, mortality risk was lower for patients intubated after a failed NIV trial only in the first 10 days of follow-up and not in the 11 to 60-day mortality. On the other side, the fact that the oldest patients and those with severe coexisting disease were treated only with oxygen carrying a higher early mortality was clearly described by the difference between the crude HRs and the HRs adjusted only be age. This finding together with the information of a mean IMV length of almost 20 days may support the idea that those patients would have not survived anyway to the IMV and, therefore, endorse the decisions taken by the multidisciplinary team. Moreover, our results suggest that initial management of severe hypoxemia by oxygen or NIV might be a valuable alternative to immediate IMV in the occurrence of limited available resources.

### Thoughts on the shared decision-making process

The decision about the best breathing support to be provided to COVID-19 patients is anything but simple. Although often severely hypoxic, they tend to present less severe dyspnea than expected, probably because many patients, at least in the early stages of the disease, have normal pulmonary compliance and therefore exert limited inspiratory efforts. In patients whose lung compliance tends to progressively decrease, the inspiratory effort increases and vigorous inspiratory effort can contribute to lung injury (Patient Self-Inflicted Lung Injury–P-SILI).^20^ This feature has been supposed to rise morbidity and mortality. Therefore, early mechanical ventilatory support has been advocated for COVID-19 associated respiratory distress.^21^ Unfortunately, criteria to intubate COVID-19 patients are controversial and the decision may locally reflect the available resources.^22^

Older age and comorbidities burden have been largely reported as the two main conditions associated with increased mortality risk,^4,8,23^ so that prioritization of younger patients has been advised in case of shortage of resources.^24^ Notably, in the present investigation –although age was higher among non-survivors than survivors in each of the four groups and each calendar year was associated with a 10% increased risk for mortality– the age difference was smaller and not statistically significant among survivors and non-survivors undergoing IMV. Moreover, the median age of patients submitted to IMV in our study (69 years) was slightly higher than reported by other authors (59 to 64 years),^2,4,8^ suggesting that the adopted criteria at our Institution were less restrictive in term of age. Furthermore, once a patient was considered as potentially salvageable by ICU admission and IMV, the length of IMV was not a criterion to withdrawing treatment. We strongly think that this ethically crucial decision could be widely considered.

Another interesting finding of our study was that similar P/F ratio was found among patients treated with NIV-only or with IMV, either preceded by a NIV trial or not. Therefore, we speculated that a low P/F ratio should not be the only criterion to decide which patient would benefit from IMV. It should be noted that among patients intubated after a NIV trial or receiving immediate IMV, the latter group had a higher SOFA score and SAPS (Table 2). These findings highlight the fundamental role played by our organizational strategy, allowing to ensure a tailored treatment to each patient by taking into account the level of care that would have more benefited for her/him, and to daily re-discuss each decision at the light of new clinical reasons or changes in the resources availability. For example, during the very early phase of the outbreak not all patients with appropriate indications were treated with NIV due to the scarce availability of helmets, while their subsequent increase in availability allowed more targeted choices in the following days.

Moreover, this strategy contributed to create a more collaborative way to approach difficult decisions, supporting healthcare professionals, especially the younger ones, during such ethically and emotionally demanding decisions.^22^

### Strength and limitations

The main strength of the study consisted in having described the impact of the SARS-CoV-2 epidemic in an entire province. Since all patients with moderate to severe COVID-19 were managed at the same hospital, a shared and homogeneous standard of care was guaranteed. Moreover, follow-up was established at 60 days, taking into account all respiratory supports and without patients’ loss. As many COVID-19 patients require prolonged IMV, a short follow-up time is an important limitation for the majority of the reports published so far.

The number of patients requiring ventilatory support was smaller compared to other studies.^2,3,10^ Consequently, the generalizability of our findings should be considered with caution.

### Conclusions

The COVID-19 outbreak has strongly challenged the healthcare systems of many countries. A multidisciplinary panel in charge of the decision of the individualized breathing approach to adopt with hospitalized COVID-19 patients maybe be a valuable option to maximize 60-day survival, dealing with the imbalance between the available resources and the clinical needs. Our findings highlight the need of high quality follow-up data that could support the decision-making for the appropriate ventilatory support strategy for COVID-19 patients.

## Data Availability

Data collected for the study appropriately deidentified, will be made available to others upon an official proposal describing the rationale and the statistical analysis plan intended to be performed by the proponent institution. A signed data access agreement will be required for handling the data.

## Acknowledgements

The Authors thank all nurses and physicians who cared for COVID-19 patients during the study period

